# Automated Phenotyping of Mitral Stenosis Using Deep Learning

**DOI:** 10.64898/2026.03.03.26347557

**Authors:** Hirotaka Ieki, Yuki Sahashi, Miloš Vukadinovic, Meenal Rawlani, Isabel Kim, Andrew P. Ambrosy, Alan S. Go, Bryan He, Paul Cheng, David Ouyang

## Abstract

**Background and Aims:** Accurate classification of mitral stenosis (MS) remains a significant clinical challenge. This study aimed to develop an artificial intelligence (AI) framework to automatically detect clinically significant MS from echocardiography.

**Methods:** We developed EchoNet-MS, an open-source end-to-end integrated approach combining video based convolutional neural networks to assess MS severity and differentiate rheumatic etiology from echocardiography and validated its performance across four cohorts.

**Results:** EchoNet-MS was trained and validated in total of 431,612 videos from 44,671 studies from three different healthcare system. Combining assessments from multiple echocardiographic videos, the model was trained on a Kaiser Permanente Northern California (KPNC) cohort of 8,677 studies from 7,576 patients with a range of MS severity. The model was validated on a KPNC held-out test cohort (N=1,623) and a temporally distinct cohort (N=19,206), as well as Stanford Healthcare (SHC) cohort (N=3,333) and Cedars-Sinai Medical Center (CSMC) cohort (N=72,909). EchoNet-MS achieved excellent discrimination of severe MS with AUC 0.937 [95% CI: 0.913 – 0.958] in the KPNC held-out cohort, 0.994 [0.986 – 0.999] in the temporally distinct cohort, 0.991 [0.986 – 0.995] in SHC, and 0.973 [0.958 – 0.987] in CSMC. The model achieved excellent performance in classifying both rheumatic or non-rheumatic MS with AUC ranging from 0.890 and 0.967.

**Conclusions:** EchoNet-MS accurately assesses MS severity and etiology using information from multiple echocardiographic views. Its strong performance generalizes robustly to external cohorts and shows potential as an automated clinical decision support tool.

## Introduction

Mitral stenosis (MS) is a clinically important valvular disease with substantial global health burden that is associated with atrial fibrillation, thromboembolic events, pulmonary hypertension, heart failure and morbidity^1,2^. Rheumatic MS, caused by prior rheumatic fever, is more prevalent in women and in low-income countries, and has been associated with a higher risk of thromboembolic complications. In contrast, non-rheumatic degenerative or calcific MS is increasingly observed in elderly patients and those with chronic kidney disease, presents distinct anatomic and procedural challenges, often complicating surgical intervention, and requiring different treatment strategies^2,3^.

Accurate diagnosis and characterization are essential for identifying patients who require intervention, tailoring monitoring frequency for disease progression, and guiding treatment decisions. Transthoracic echocardiography (TTE) is the primary modality for MS assessment, providing both severity and morphological evaluation^4^. However, comprehensive interpretation often requires integration of multiple views, can be time-consuming, and may vary across readers—particularly in high-throughput clinical practice^5^. With continuing advances in percutaneous mitral intervention, accurate assessment of MS is critical for timely referral and treatment^6,7^.

Recent advances in deep learning methods have enabled automated severity assessment of valvular heart disease, including mitral regurgitation (MR), aortic regurgitation (AR), tricuspid regurgitation (TR) and aortic stenosis (AS) from echocardiography images or videos^8–13^, enabling a high throughput reproducible phenotyping. More recently, open-source multi-task foundation models have been proposed to support broad phenotyping at scale, demonstrating strong performance across multiple labels including MS^14,15^, yet robust, externally validated methods specific for MS detection and etiologic characterization remain limited. Here, we developed a first open-source deep learning framework, EchoNet-MS, which integrates information from multiple echocardiographic views and Doppler images to assess the severity and etiology of MS and validated the framework across multiple institutions to demonstrate its generalizability.

## Methods

### Study population, imaging and data source

In this multicenter retrospective study, we used transthoracic echocardiography (TTE) data from three different medical centers: Kaiser Permanente Northern California (KPNC; California, USA), Stanford Healthcare (SHC; Stanford California, USA), and Cedars-Sinai Medical Center (CSMC; Los Angeles, California, USA) (**Figure 1**). In KPNC, 10,300 studies conducted between January 2022 and December 2024 were sampled, with cases without MS downsampled to mitigate class imbalance during model training. Patients who had mitral valve surgery before TTE were excluded. This cohort was split into a derivation dataset (training + validation; 86612 videos from 8,677 TTE studies from 7,576 patients) and a held-out test cohort (16008 videos from 1,623 TTE studies from 1,408 patients), ensuring no patient overlap between the subsets. The final derivation dataset comprised of 8,677 studies from 7576 patients with 86,612 echocardiographic videos. To assess temporal generalizability, we additionally collected 34,371 studies from 33,534 conducted between January and August 2025. All the videos were recorded from adult participants on Philips (Amsterdam, The Netherlands) EPIQ 7C, EPIQ CVx, Affiniti 70C or Affiniti CVx machines. The acquisition of images followed a comprehensive and standardized protocol in-line with current recommendations^4,16,17^.

**Figure 1.**
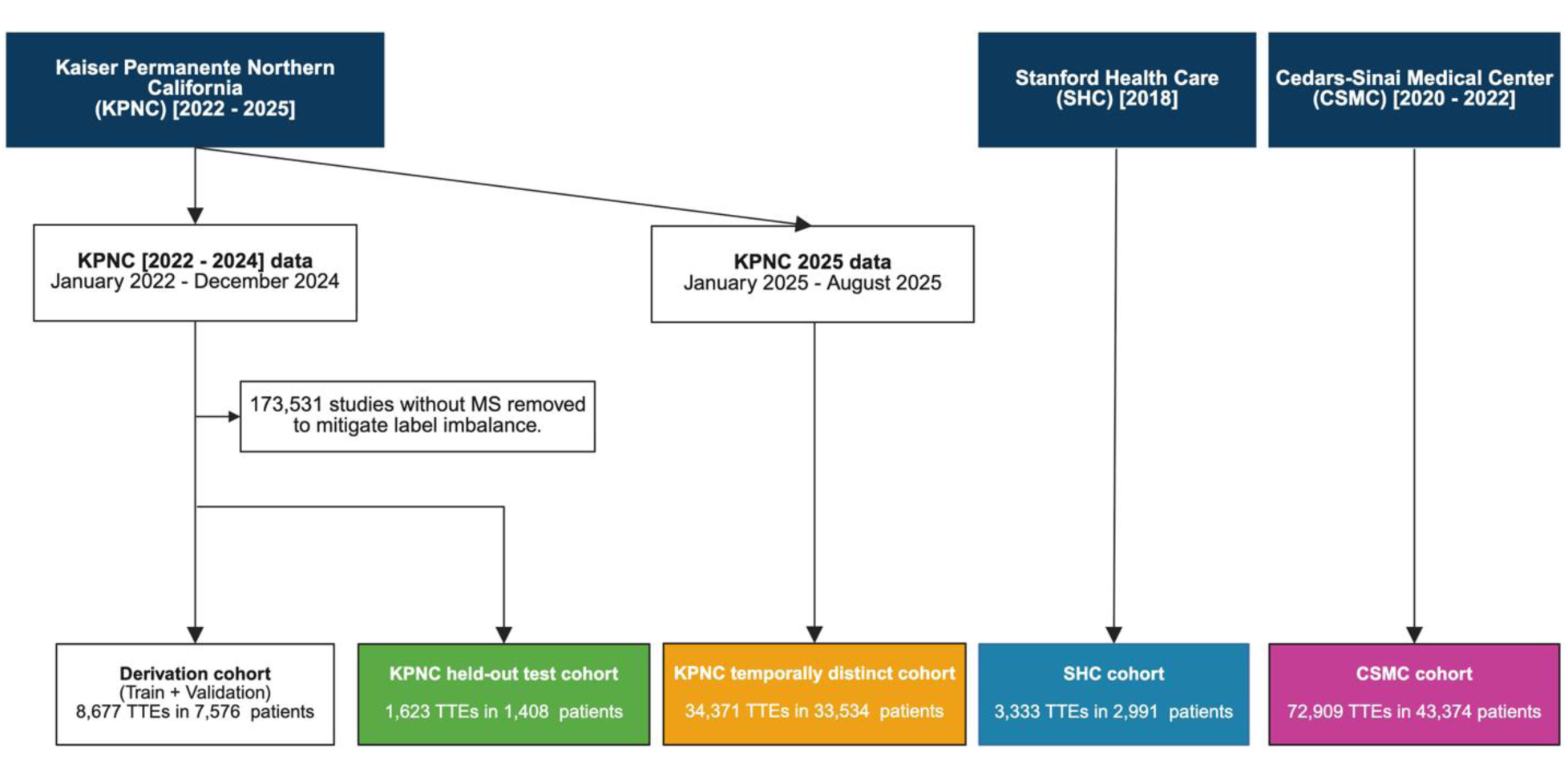
Data flowchart. The construction of the study cohorts from echocardiographic databases from three different medical centers: Kaiser Permanente Northern California (KPNC), Stanford Health Care (SHC), and Cedars-Sinai Medical Center (CSMC). From the KPNC database, 10,300 TTEs from 8,984 patients (2022–2024) were filtered to form a balanced cohort by removing 17,531 studies without MS. This cohort was split into a derivation dataset (8,677 TTEs for training and validation data) and a held-out test cohort (1,623 TTEs). Separately, 34,371 TTEs conducted between January and August 2025 were used as the temporally distinct test cohort. There is no patient overlap between the cohorts. From the Stanford and Cedars-Sinai databases, external validation cohorts were formed with 3,333 TTEs from 2,991 patients and 72,909 TTEs from 43,374 patients, respectively. Created in https://BioRender.com

Additionally, our model was evaluated on two distinct external cohorts from separate healthcare systems. A total of 3,333 studies from 2,991 patients conducted between 2016 and 2018 were extracted from SHC database. In addition, 72,909 TTEs from 43,377 patients conducted between 2020 and 2022 at CSMC were extracted. The SHC and CSMC echocardiography lab uses Philips EPIQ 7 or IE33 machines and follows a similar standardized comprehensive protocol. MS severity was extracted from finalized echocardiography report. For all cohorts, MS severity was categorized as ‘no, ‘mild’, mild to moderate’, ‘moderate’, ‘moderate to severe’ or ‘severe’. For model training and evaluation, intermediate MS categories (e.g. “mild to moderate”) were upgraded to the upper categories (e.g. “moderate”). No additional adjudication or consensus review was performed as part of this study, reflecting real-world reporting practice.

### AI model development

We developed a two-stage framework for MS assessment, termed EchoNet-MS, consisting of deep learning predictions from videos and ensemble integration across different views (**Figure 2**). In the first stage, four separate video-based convolutional neural networks (CNNs) based on the ResNet R(2+1)D architecture^18^ were trained to classify MS severity from standard echocardiographic views: parasternal long axis (PLAX), apical four chamber views (A4C), PLAX color Doppler, A4C color Doppler. Additionally, an image-based CNN (ResNet50 architecture^19^) were trained to classify MS severity and to predict mean mitral valve inflow gradient from mitral valve continuous wave Doppler images. The images were stored as Digital Imaging and Communications in Medicine (DICOM) files, and these files underwent pre-processing to ensure consistency and extraneous details beyond the ultrasound section were removed, and metadata was de-identified. The input videos and images were processed to mask the embedded texts and were resized to 112 × 112 and 224 × 224 pixels, respectively. The videos were subsampled by one in every two frames to yield 16 frames. Cross-entropy loss for classification or mean squared error for regression were employed as the loss function. Training was performed on NVIDIA L40S GPUs for up to 100 epochs, with early stopping after 10 epoch based on validation area under the receiver operating characteristic curve (AUROC) or mean absolute error (MAE) to prevent overfitting. All the models were trained using the Pytorch 2.5.0 and PyTorch Lightning 2.5.1 ^20^ deep learning framework with the Rectified Adam Schedule Free optimizer ^21^ with a batch size of 128 and mixed-precision training (bfloat16) ^22^.

**Figure 2.**
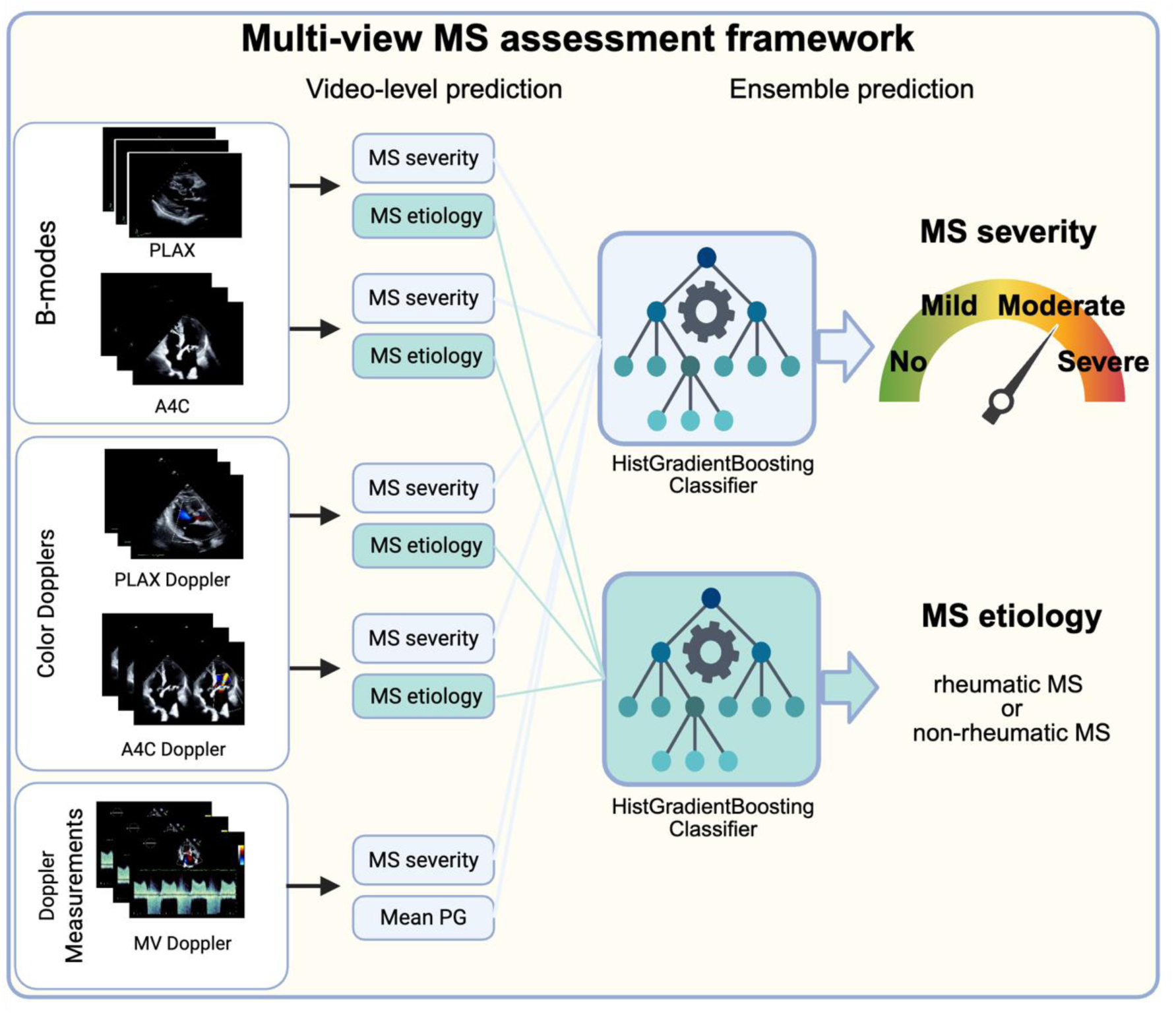
Overview of the model. In the EchoNet-MS framework, we trained video-based convolutional neural network (CNN) models to predict the severity of mitral stenosis (MS) from four distinct transthoracic echocardiographic views including both B-modes [parasternal long axis (PLAX) and apical four chamber (A4C) views] and Color dopplers [PLAX Doppler, A4C Doppler] and mitral valve inflow Doppler image. These outputs were integrated through a tree-based ensemble approach to generate a final prediction of MS severity. In parallel, video-based CNN models were trained to classify MS etiology (rheumatic vs. non-rheumatic degenerative MS). These outputs were similarly combined through an ensemble model to produce the final etiology prediction. Created in https://BioRender.com

The second stage of the EchoNet-MS consisted of an ensemble model. The outputs from the six networks were used as input features for a machine learning ensemble implemented with the HistGradientBoostingClassifier in scikit-learn^23^. The final output is the probabilities of four MS severity classes (‘no’, ‘mild’, ‘moderate’ or ‘severe’). When multiple videos were available for the same view, the model outputs were averaged prior to integration. The ensemble classifier is robust to missing values, thereby allowing inclusion of studies that did not contain all views.

Additionally, a video-based CNNs were trained to classify rheumatic or non-rheumatic MS. Using the same data splits as the MS severity model, patients with at least mild MS were identified, and the presence or absence of rheumatic changes was extracted from the echocardiography reports to serve as ground-truth labels. Four view specific video-based CNN (PLAX, A4C, PLAX color Doppler and A4C color Doppler) were trained for rheumatic classification following the same preprocessing procedures, architecture, and training strategy as described above. Model outputs from the individual networks were subsequently integrated using a HistGradientBoostingClassifier ensemble.

### Statistical analysis

The EchoNet-MS framework was evaluated across four independent test cohorts. For the description of the baseline cohort characteristics, categorical variables are presented as counts and percentages, while continuous variables are expressed as mean and standard deviation. We calculated the area under the receiver operating characteristic curve (AUROC) as the main outcome. Area under the precision-recall curve (AUPRC), positive predictive value (PPV), negative predictive value (NPV), sensitivity, specificity and F1 score were calculated for clinically significant MS, which was defined as greater than moderate MS and severe MS. All 95% confidence intervals were calculated from 10,000 bootstrapping replicates. Subgroup analysis was conducted in KPNC temporally distinct cohort and CSMC cohort to assess model performance in patients with different ranges of age, sex, body mass index (BMI), left ventricular ejection fraction (LVEF), associated comorbidities (at least moderate AS, MR, and TR), and other clinical characteristics. To compare the performance relative to recent open-source foundation models, we performed comparisons against publicly available models (EchoPrime^14^ and PanEcho^15^) on the KPNC held-out and temporally distinct cohorts. To understand which regions of the echocardiographic videos contributed most for AI model assessment, we employed smooth grad saliency mapping^24^. To characterize misclassified cases, we assessed the discrepancy between predicted and ground-truth MS severity and compared clinical characteristics between misclassified and correctly classified cases using KPNC temporally distinct cohort. Analyses were performed using Python (version 3.12.3) and R (version 4.5.1)

## Results

### Study population

A total of 1,294,979 images and videos from 112,236 studies, obtained from 81,300 unique patients from KPNC, SHC and CSMC, were used for model development and evaluation in this study. The KPNC derivation and held-out test cohort patients were older and included a higher proportion of female patients (mean age (years); male (%): 72.3 ± 15.3; 37.8% in derivation cohort, 72.0 ± 15.0; 39.6% in held-out test cohort) compared to the other test cohort (65.3 ± 16.9; 49.3%, in KPNC temporally distinct test cohort, 65.7 ± 16.2; 51.5% in the SHC cohort, and 65,7 ± 16.8; 56.0% in the CSMC cohort), as the KPNC derivation and held-out test cohort were enriched for MS patients to mitigate class imbalance. The proportion, racial/ethnic groups, the mean left ventricular ejection fraction, and major comorbidities were similar across datasets. A detailed description of the characteristics of the cohort is shown in **Table 1**.

**Table 1.**
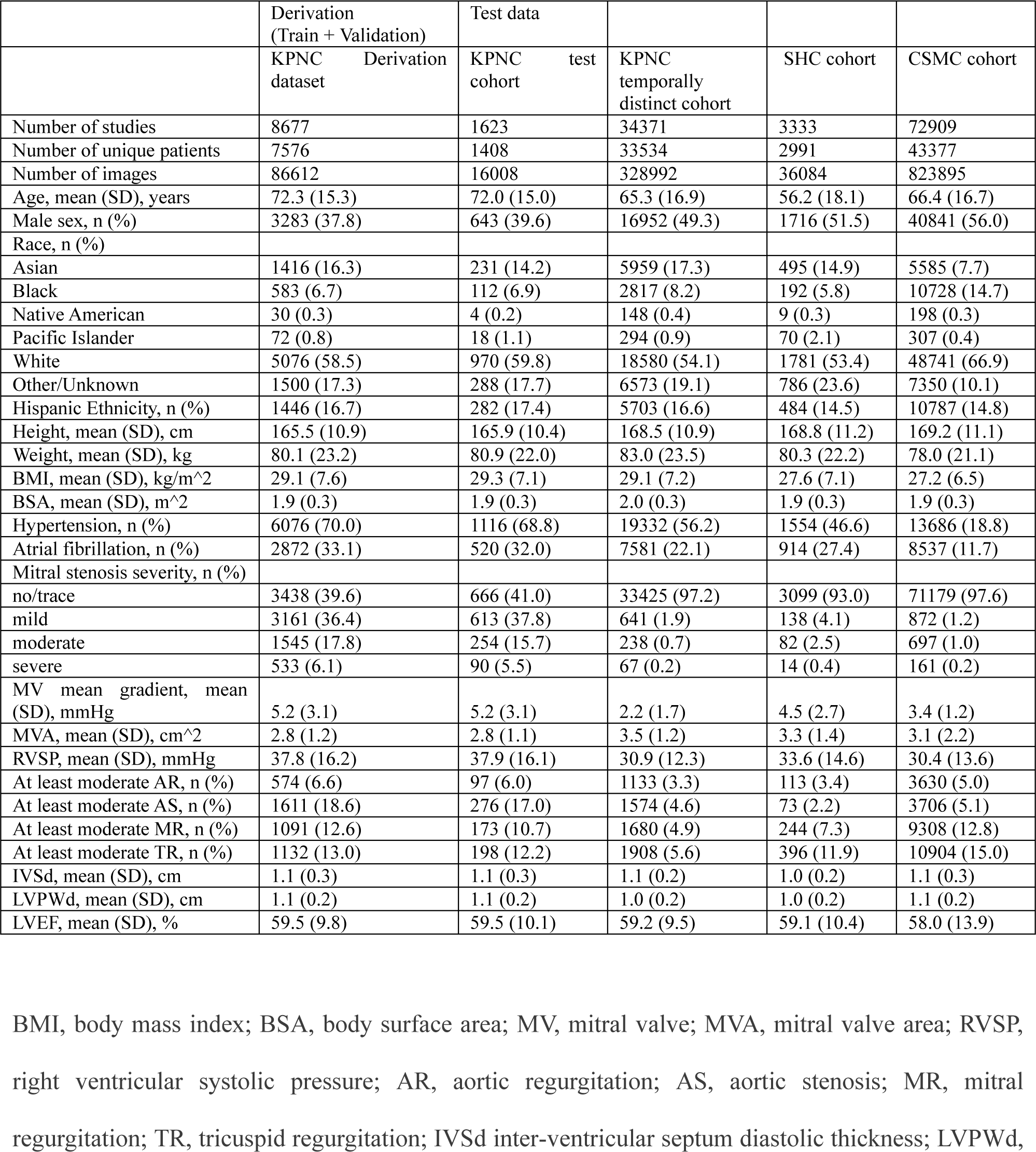
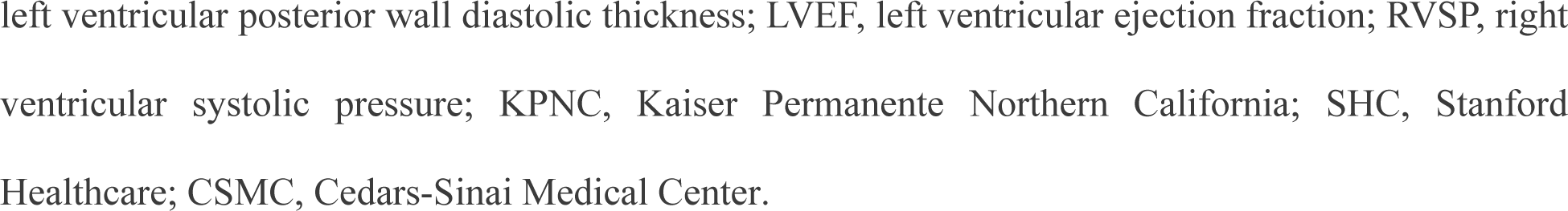
Table of baseline demographic and echocardiographic characteristics in each cohort.

|  | Derivation<br>(Train + Validation) | Test data |  |  |  |
| --- | --- | --- | --- | --- | --- |
|  | KPNC Derivation<br>dataset | KPNC test<br>cohort | KPNC<br>temporally<br>distinct cohort | SHC cohort | CSMC cohort |
| Number of studies | 8677 | 1623 | 34371 | 3333 | 72909 |
| Number of unique patients | 7576 | 1408 | 33534 | 2991 | 43377 |
| Number of images | 86612 | 16008 | 328992 | 36084 | 823895 |
| Age, mean (SD), years | 72.3 (15.3) | 72.0 (15.0) | 65.3 (16.9) | 56.2 (18.1) | 66.4 (16.7) |
| Male sex, n (%) | 3283 (37.8) | 643 (39.6) | 16952 (49.3) | 1716 (51.5) | 40841 (56.0) |
| Race, n (%) |  |  |  |  |  |
| Asian | 1416 (16.3) | 231 (14.2) | 5959 (17.3) | 495 (14.9) | 5585 (7.7) |
| Black | 583 (6.7) | 112 (6.9) | 2817 (8.2) | 192 (5.8) | 10728 (14.7) |
| Native American | 30 (0.3) | 4 (0.2) | 148 (0.4) | 9 (0.3) | 198 (0.3) |
| Pacific Islander | 72 (0.8) | 18 (1.1) | 294 (0.9) | 70 (2.1) | 307 (0.4) |
| White | 5076 (58.5) | 970 (59.8) | 18580 (54.1) | 1781 (53.4) | 48741 (66.9) |
| Other/Unknown | 1500 (17.3) | 288 (17.7) | 6573 (19.1) | 786 (23.6) | 7350 (10.1) |
| Hispanic Ethnicity, n (%) | 1446 (16.7) | 282 (17.4) | 5703 (16.6) | 484 (14.5) | 10787 (14.8) |
| Height, mean (SD), cm | 165.5 (10.9) | 165.9 (10.4) | 168.5 (10.9) | 168.8 (11.2) | 169.2 (11.1) |
| Weight, mean (SD), kg | 80.1 (23.2) | 80.9 (22.0) | 83.0 (23.5) | 80.3 (22.2) | 78.0 (21.1) |
| BMI, mean (SD), kg/m<sup>2</sup> | 29.1 (7.6) | 29.3 (7.1) | 29.1 (7.2) | 27.6 (7.1) | 27.2 (6.5) |
| BSA, mean (SD), m<sup>2</sup> | 1.9 (0.3) | 1.9 (0.3) | 2.0 (0.3) | 1.9 (0.3) | 1.9 (0.3) |
| Hypertension, n (%) | 6076 (70.0) | 1116 (68.8) | 19332 (56.2) | 1554 (46.6) | 13686 (18.8) |
| Atrial fibrillation, n (%) | 2872 (33.1) | 520 (32.0) | 7581 (22.1) | 914 (27.4) | 8537 (11.7) |
| Mitral stenosis severity, n (%) |  |  |  |  |  |
| no/trace | 3438 (39.6) | 666 (41.0) | 33425 (97.2) | 3099 (93.0) | 71179 (97.6) |
| mild | 3161 (36.4) | 613 (37.8) | 641 (1.9) | 138 (4.1) | 872 (1.2) |
| moderate | 1545 (17.8) | 254 (15.7) | 238 (0.7) | 82 (2.5) | 697 (1.0) |
| severe | 533 (6.1) | 90 (5.5) | 67 (0.2) | 14 (0.4) | 161 (0.2) |
| MV mean gradient, mean (SD), mmHg | 5.2 (3.1) | 5.2 (3.1) | 2.2 (1.7) | 4.5 (2.7) | 3.4 (1.2) |
| MVA, mean (SD), cm<sup>2</sup> | 2.8 (1.2) | 2.8 (1.1) | 3.5 (1.2) | 3.3 (1.4) | 3.1 (2.2) |
| RVSP, mean (SD), mmHg | 37.8 (16.2) | 37.9 (16.1) | 30.9 (12.3) | 33.6 (14.6) | 30.4 (13.6) |
| At least moderate AR, n (%) | 574 (6.6) | 97 (6.0) | 1133 (3.3) | 113 (3.4) | 3630 (5.0) |
| At least moderate AS, n (%) | 1611 (18.6) | 276 (17.0) | 1574 (4.6) | 73 (2.2) | 3706 (5.1) |
| At least moderate MR, n (%) | 1091 (12.6) | 173 (10.7) | 1680 (4.9) | 244 (7.3) | 9308 (12.8) |
| At least moderate TR, n (%) | 1132 (13.0) | 198 (12.2) | 1908 (5.6) | 396 (11.9) | 10904 (15.0) |
| IVSd, mean (SD), cm | 1.1 (0.3) | 1.1 (0.3) | 1.1 (0.2) | 1.0 (0.2) | 1.1 (0.3) |
| LVPWd, mean (SD), cm | 1.1 (0.2) | 1.1 (0.2) | 1.0 (0.2) | 1.0 (0.2) | 1.1 (0.2) |
| LVEF, mean (SD), % | 59.5 (9.8) | 59.5 (10.1) | 59.2 (9.5) | 59.1 (10.4) | 58.0 (13.9) |

BMI, body mass index; BSA, body surface area; MV, mitral valve; MVA, mitral valve area; RVSP,
right ventricular systolic pressure; AR, aortic regurgitation; AS, aortic stenosis; MR, mitral
regurgitation; TR, tricuspid regurgitation; IVSd inter-ventricular septum diastolic thickness; LVPWd,
left ventricular posterior wall diastolic thickness; LVEF, left ventricular ejection fraction; RVSP, right ventricular systolic pressure; KPNC, Kaiser Permanente Northern California; SHC, Stanford Healthcare; CSMC, Cedars-Sinai Medical Center.

### Model performance

Our EchoNet-MS showed strong performance in distinguishing MS severity and identifying clinically significant MS, with an AUC of 0.937 [0.913 - 0.958] for detecting severe MS and 0.912 [0.896-0.927] for moderate or severe MS in the KPNC held-out test cohort, and 0.994 [0.986 - 0.999] for severe MS and 0.987 [0.984 - 0.990] for moderate or severe MS in the KPNC temporally distinct cohort (**Figure 3A**, **Figure 4A, B**). Across both KPNC cohorts, the model demonstrated consistently high NPV (KPNC held-out; 0.899 [0.883–0.915] for moderate or severe MS and 0.971 [0.962–0.979] for severe MS, KPNC temporally distinct; 0.996 [0.995–0.997] and 0.999 [0.999–0.999], respectively) (**Table 2**). We further evaluated model generalizability in two external validation cohorts. In the SHC cohort, the model achieved AUCs of 0.959 [0.940–0.975] for moderate or severe MS and 0.991 [0.986–0.995] for severe MS. In the CSMC cohort, performance remained robust, with AUCs of 0.969 [0.964–0.974] for moderate or severe MS and 0.973 [0.958–0.987] for severe MS (**Figure 3A**, **Figure 4C, D**). Together, these results demonstrate consistent discrimination across independent healthcare systems, supporting the generalizability of the model. Although the PPV varies across cohorts (0.333–0.827), likely reflecting the low prevalence of severe MS, NPV for detecting severe MS and moderate or severe MS remained consistently high across all cohorts (0.899 – 0.999), indicating that the model can reliably exclude clinically significant MS.

**Figure 3.**
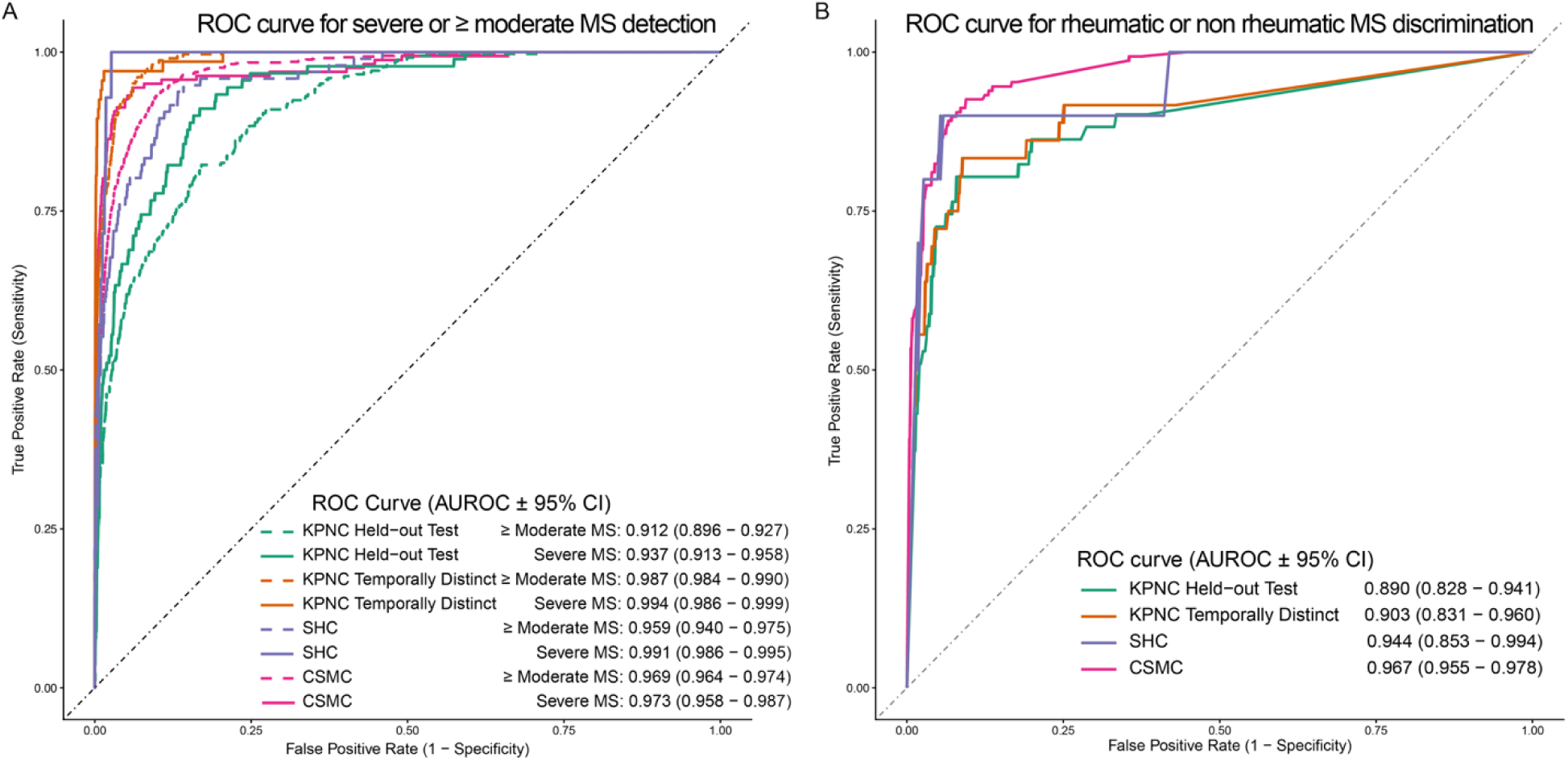
(A) Receiver operating characteristic (ROC) curves for detection of severe or at least moderate mitral stenosis (MS). (B) ROC curves for MS etiology classification. Results in Kaiser Permanente Northern California (KPNC) held-out test cohort (green), KPNC temporally distinct cohort (orange), Stanford Healthcare (SHC) cohort (purple), and Cedars-Sinai Medical Center (CSMC) cohort (pink) are shown. Predictions for severe and at least moderate are shown in solid and dash lines, respectively. AUROC area under the receiver operating characteristic curve; CI, confidence interval.

**Figure 4.**
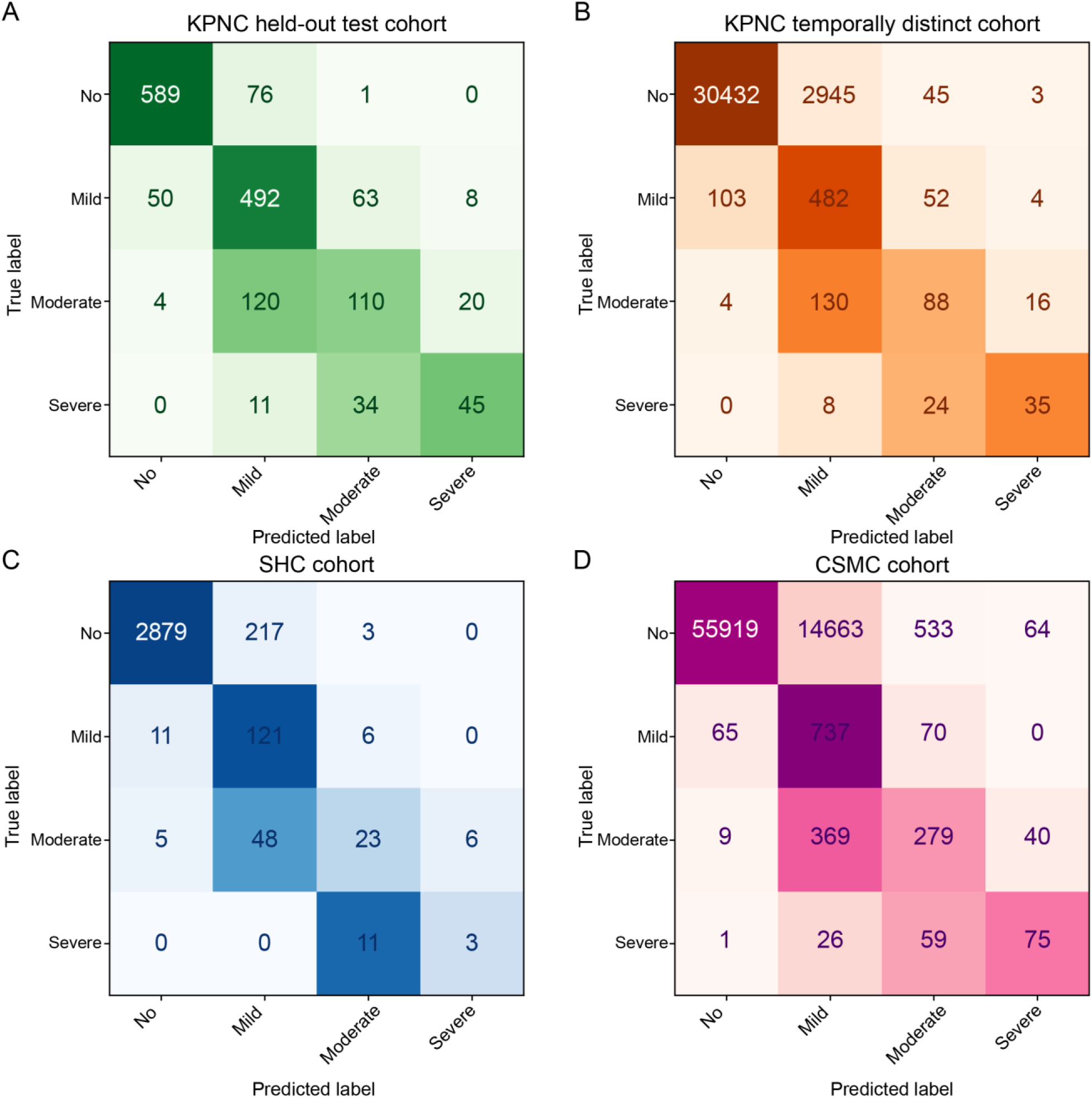
Confusion matrix of MS severity classification on KPNC test cohort (A), KPNC temporally distinct cohort (B), SHC cohort (C), and CSMC cohort (D). Confusion matrix colormap values were scaled based on the proportion of actual disease cases in each class that were predicted in each possible disease category.

**Table 2.**
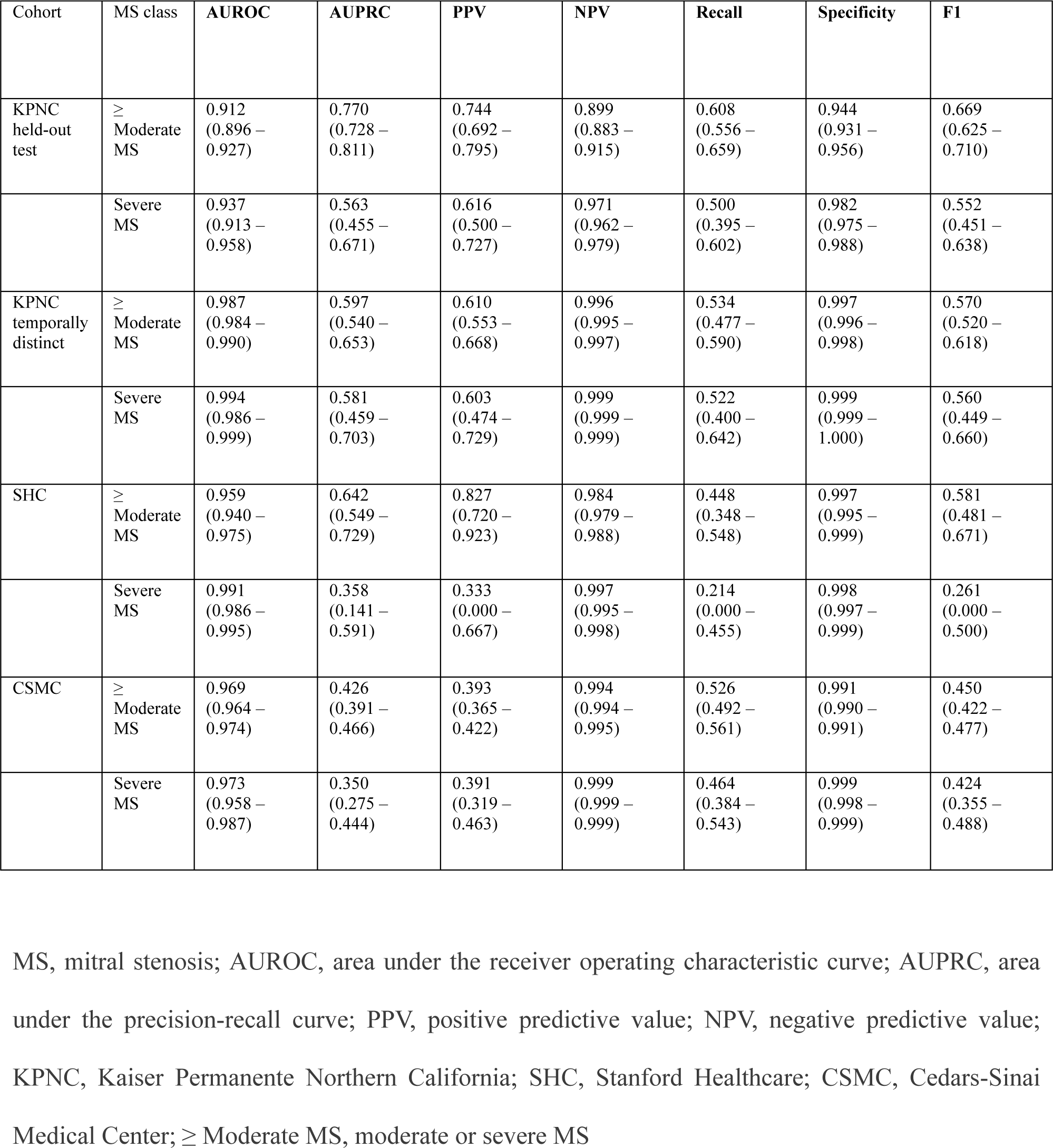
Different performance metrics of EchoNet-MS to detect severe or at least moderate mitral stenosis in the test cohorts.

| Cohort | MS class | AUROC | AUPRC | PPV | NPV | Recall | Specificity | F1 |
| --- | --- | --- | --- | --- | --- | --- | --- | --- |
| KPNC held-out test | ≥ Moderate MS | 0.912<br>(0.896 – 0.927) | 0.770<br>(0.728 – 0.811) | 0.744<br>(0.692 – 0.795) | 0.899<br>(0.883 – 0.915) | 0.608<br>(0.556 – 0.659) | 0.944<br>(0.931 – 0.956) | 0.669<br>(0.625 – 0.710) |
|  | Severe MS | 0.937<br>(0.913 – 0.958) | 0.563<br>(0.455 – 0.671) | 0.616<br>(0.500 – 0.727) | 0.971<br>(0.962 – 0.979) | 0.500<br>(0.395 – 0.602) | 0.982<br>(0.975 – 0.988) | 0.552<br>(0.451 – 0.638) |
| KPNC temporally distinct | ≥ Moderate MS | 0.987<br>(0.984 – 0.990) | 0.597<br>(0.540 – 0.653) | 0.610<br>(0.553 – 0.668) | 0.996<br>(0.995 – 0.997) | 0.534<br>(0.477 – 0.590) | 0.997<br>(0.996 – 0.998) | 0.570<br>(0.520 – 0.618) |
|  | Severe MS | 0.994<br>(0.986 – 0.999) | 0.581<br>(0.459 – 0.703) | 0.603<br>(0.474 – 0.729) | 0.999<br>(0.999 – 0.999) | 0.522<br>(0.400 – 0.642) | 0.999<br>(0.999 – 1.000) | 0.560<br>(0.449 – 0.660) |
| SHC | ≥ Moderate MS | 0.959<br>(0.940 – 0.975) | 0.642<br>(0.549 – 0.729) | 0.827<br>(0.720 – 0.923) | 0.984<br>(0.979 – 0.988) | 0.448<br>(0.348 – 0.548) | 0.997<br>(0.995 – 0.999) | 0.581<br>(0.481 – 0.671) |
|  | Severe MS | 0.991<br>(0.986 – 0.995) | 0.358<br>(0.141 – 0.591) | 0.333<br>(0.000 – 0.667) | 0.997<br>(0.995 – 0.998) | 0.214<br>(0.000 – 0.455) | 0.998<br>(0.997 – 0.999) | 0.261<br>(0.000 – 0.500) |
| CSMC | ≥ Moderate MS | 0.969<br>(0.964 – 0.974) | 0.426<br>(0.391 – 0.466) | 0.393<br>(0.365 – 0.422) | 0.994<br>(0.994 – 0.995) | 0.526<br>(0.492 – 0.561) | 0.991<br>(0.990 – 0.991) | 0.450<br>(0.422 – 0.477) |
|  | Severe MS | 0.973<br>(0.958 – 0.987) | 0.350<br>(0.275 – 0.444) | 0.391<br>(0.319 – 0.463) | 0.999<br>(0.999 – 0.999) | 0.464<br>(0.384 – 0.543) | 0.999<br>(0.998 – 0.999) | 0.424<br>(0.355 – 0.488) |

MS, mitral stenosis; AUROC, area under the receiver operating characteristic curve; AUPRC, area under the precision-recall curve; PPV, positive predictive value; NPV, negative predictive value; KPNC, Kaiser Permanente Northern California; SHC, Stanford Healthcare; CSMC, Cedars-Sinai Medical Center; ≥ Moderate MS, moderate or severe MS

### Subgroup analysis

Analyzing patient subgroups further supported the generalizability of the model (**Table 3**). In the KPNC temporally distinct cohort, older patients (moderate or severe MS/severe MS: AUROC 0.983 [95% CI, 0.979–0.986] / 0.993 [0.983–0.999]), males (0.987 [0.982–0.992] / 0.987 [0.958–1.000]), females (0.985 [0.981–0.989] / 0.995 [0.990–0.999]), patients with obesity (BMI > 35: 0.986 [0.978–0.992] / 0.987 [0.960–1.000]), those with LVEF ≤ 50% (0.983 [0.975–0.991] / 0.997 [0.990–1.000]), and LVEF > 50% (0.988 [0.985–0.991] / 0.994 [0.985–0.999]) all demonstrated high performance. Performance was also robust in those with AF or concomitant valvular disease such as AS, MR, TR. Similar results were observed in the CSMC cohort: older patients (0.962 [0.954–0.968] / 0.971 [0.949–0.989]), males (0.964 [0.950–0.976] / 0.976 [0.935–0.997]), females (0.966 [0.959–0.973] / 0.975 [0.956–0.990]), obese patients (0.967 [0.952–0.980] / 0.997 [0.993–0.999]), those with LVEF ≤ 50% (0.955 [0.924–0.977] / 0.996 [0.992–0.999]), and LVEF > 50% (0.973 [0.966–0.978] / 0.981 [0.966–0.993]) consistently achieved high AUCs. Across both cohorts, severe MS detection generally outperformed moderate or severe MS detection, suggesting greater discriminative ability for severe cases.

**Table 3.** EchoNet-MS subgroup analysis.

| Subset | KPNC temporally distinct | AUROC |  | CSMC | AUROC |  |
| --- | --- | --- | --- | --- | --- | --- |
|  | Studies, No. | ≥ Moderate MS | Severe MS | Studies, No. | ≥ Moderate MS | Severe MS |
| Age > 60 | 23079 | 0.983 (0.979 – 0.986) | 0.993 (0.983 – 0.999) | 49214 | 0.962 (0.954 – 0.968) | 0.971 (0.949 – 0.989) |
| Age ≤ 60 | 11284 | 0.995 (0.987 – 0.999) | 0.995 (0.985 – 1.000) | 23695 | 0.977 (0.950 – 0.992) | 0.992 (0.979 – 0.999) |
| Male | 16947 | 0.987 (0.982 – 0.992) | 0.987 (0.958 – 1.000) | 40841 | 0.964 (0.950 – 0.976) | 0.976 (0.935 – 0.997) |
| Female | 17401 | 0.985 (0.981 – 0.989) | 0.995 (0.990 – 0.999) | 32061 | 0.966 (0.959 – 0.973) | 0.975 (0.956 – 0.990) |
| White | 18580 | 0.986 (0.982 – 0.989) | 0.993 (0.980 – 0.999) | 48741 | 0.963 (0.954 – 0.971) | 0.969 (0.941 – 0.990) |
| Other race | 15791 | 0.988 (0.983 – 0.992) | 0.995 (0.987 – 0.999) | 24168 | 0.977 (0.969 – 0.983) | 0.989 (0.980 – 0.995) |
| BMI > 35 | 5693 | 0.986 (0.978 – 0.992) | 0.987 (0.960 – 1.000) | 6607 | 0.967 (0.952 – 0.980) | 0.997 (0.993 – 0.999) |
| BMI ≤ 35 | 28664 | 0.987 (0.984 – 0.990) | 0.995 (0.987 – 0.999) | 56240 | 0.969 (0.961 – 0.976) | 0.977 (0.957 – 0.992) |
| AS | 1574 | 0.945 (0.925 – 0.962) | 0.994 (0.988 – 0.998) | 3706 | 0.909 (0.886 – 0.930) | 0.963 (0.929 – 0.989) |
| MR | 1680 | 0.973 (0.960 – 0.984) | 0.980 (0.948 – 0.998) | 9308 | 0.946 (0.933 – 0.957) | 0.967 (0.941 – 0.986) |
| TR | 1908 | 0.979 (0.969 – 0.988) | 0.997 (0.994 – 0.999) | 10904 | 0.957 (0.946 – 0.966) | 0.972 (0.949 – 0.987) |
| LVEF > 50 | 29948 | 0.988 (0.985 – 0.991) | 0.994 (0.985 – 0.999) | 57684 | 0.973 (0.966 – 0.978) | 0.981 (0.966 – 0.993) |
| LVEF ≤ 50 | 4312 | 0.983 (0.975 – 0.991) | 0.997 (0.990 – 1.000) | 14063 | 0.955 (0.924 – 0.977) | 0.996 (0.992 – 0.999) |
| AF | 7581 | 0.979 (0.972 – 0.986) | 0.996 (0.994 – 0.998) | 8537 | 0.955 (0.932 – 0.973) | 0.987 (0.976 – 0.994) |
| No AF | 26790 | 0.989 (0.986 – 0.992) | 0.991 (0.978 – 0.999) | 64372 | 0.970 (0.963 – 0.975) | 0.973 (0.950 – 0.990) |

BMI, body mass index; AS, patients with at least moderate aortic stenosis; MR, patients with at least
moderate mitral regurgitation; TR, patients with at least moderate tricuspid regurgitation; MS,
patients with at least moderate mitral stenosis; LVEF, left ventricular ejection fraction; AF, atrial
fibrillation; KPNC, Kaiser Permanente Northern California; CSMC, Cedars-Sinai Medical Center

### Explainability analysis / misclassified case analysis

Saliency map analysis revealed that the view-specific models predominantly highlighted the mitral valve and surrounding regions on two-dimensional images, as well as the corresponding color Doppler signals around the mitral valve (**Figure 5**). Among misclassifications, the vast majority (93.8% [KPNC held-out test], 98.0% [KPNC temporally distinct], 94.9% [CSMC], 97.3% [SHC], **Figure 4**) was a one-category difference. Patients with misclassified predictions were older (77.2 ± 12.9 vs. 65.3 ± 16.9 years, p < 0.001), and more prevalent in women (Female 50.6% vs 71.9%, p=0.021), while no significant difference in LVEF (59.2% vs 60.2%, p = 0.448).

**Figure 5.**
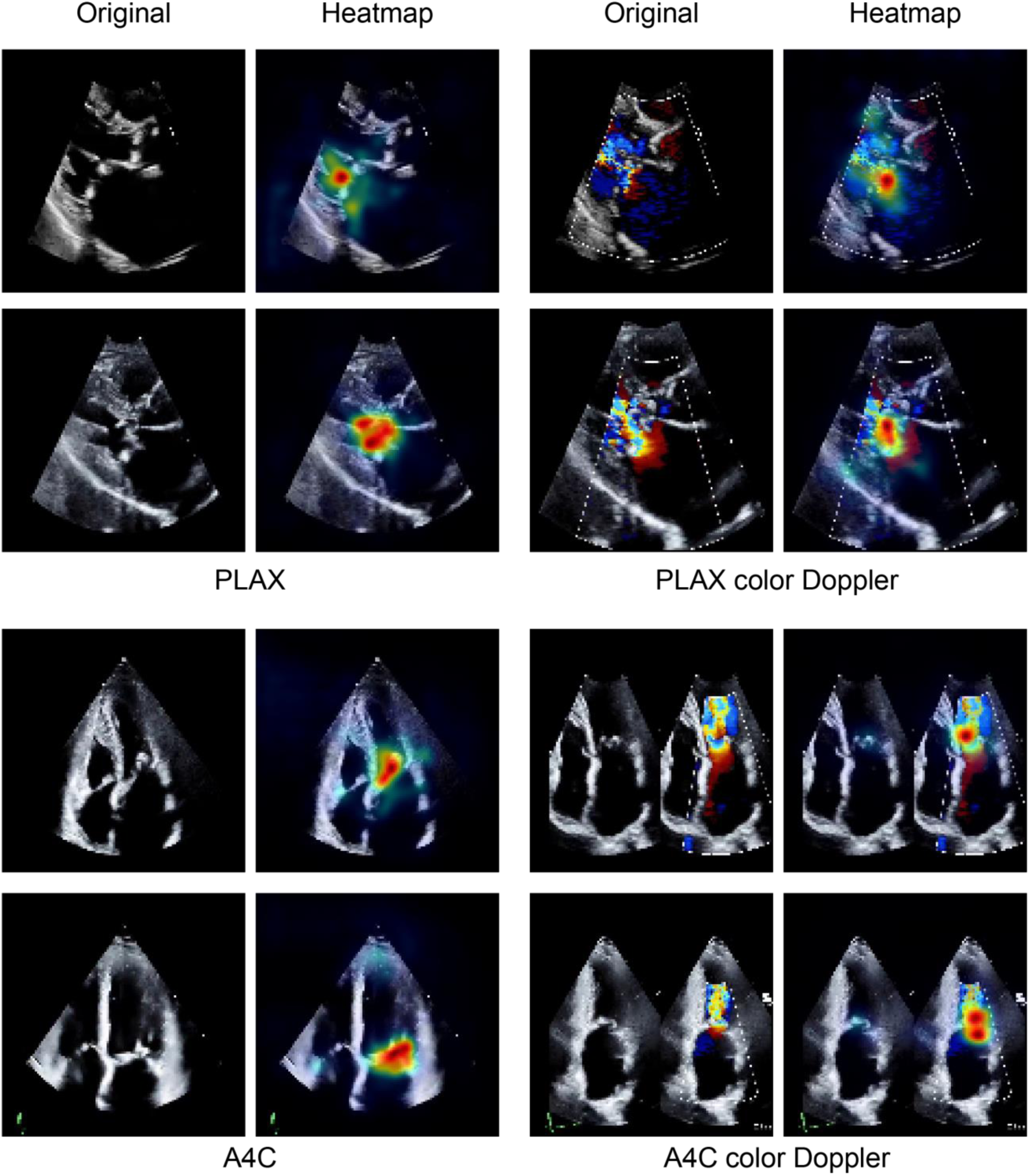
Model interpretability analysis. A. Representative Smooth Grad visualizations for each of the four echocardiographic views (parasternal long axis [PLAX], apical four chamber [A4C], and their corresponding color Doppler views) illustrating regions contributing most to model predictions of mitral stenosis (MS) severity.

### Comparison with existing models

We compared EchoNet-MS performance with the recent multi-task foundation models, EchoPrime^14^ and PanEcho^15^ on the KPNC held-out and temporally distinct cohorts. On the held-out test cohort, EchoNet-MS demonstrated higher AUROC than EchoPrime (0.937 vs. 0.846, p < 0.001) and PanEcho (0.827, p < 0.001). On the temporally distinct cohort, EchoNet-AS similarly outperformed EchoPrime (0.994 vs. 0.979, p = 0.002) and PanEcho (0.981, p = 0.011). Detailed metrics are summarized in **Table 4**.

**Table 4.**
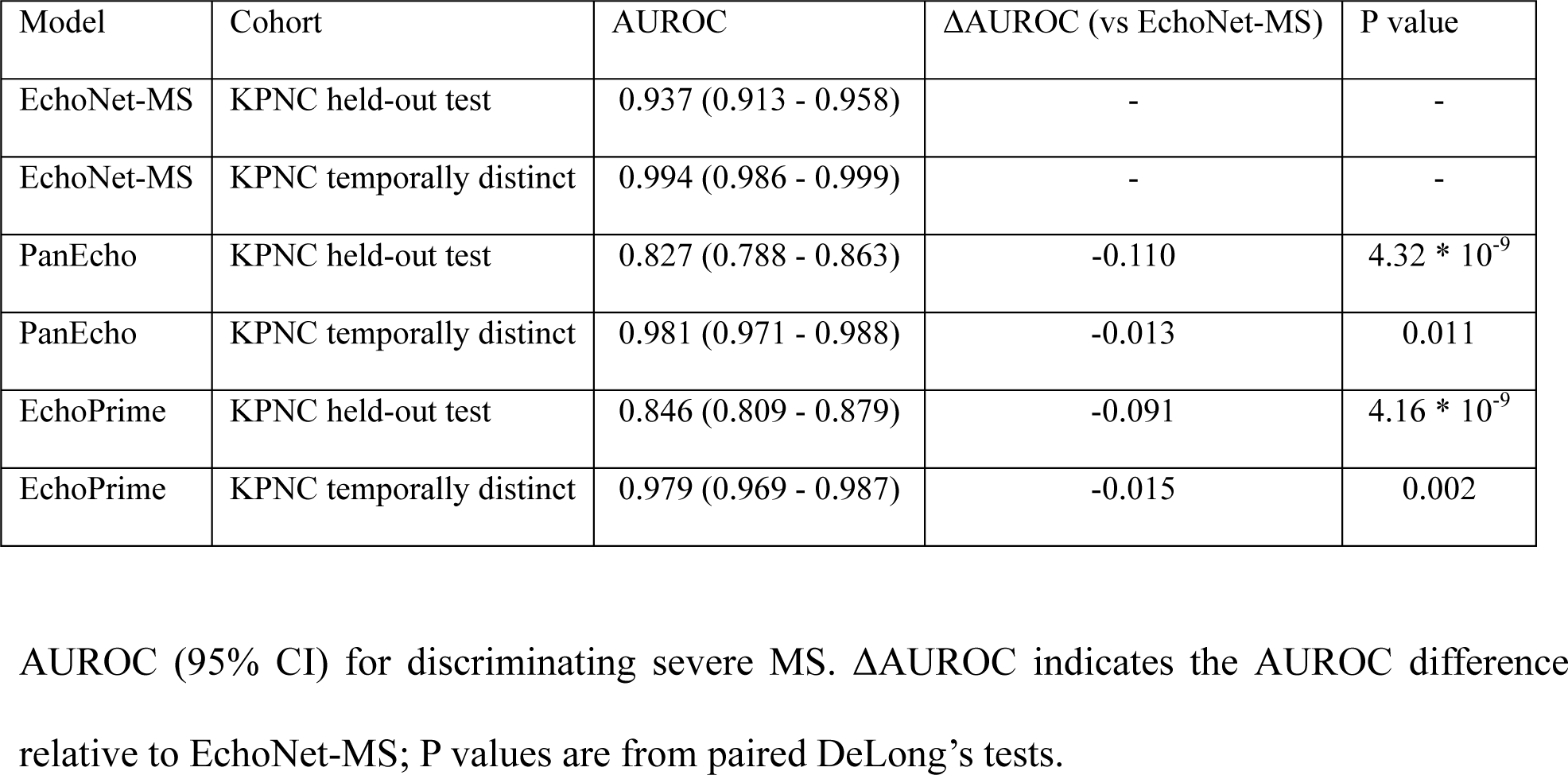
Comparison of model discrimination for severe MS.

### Rheumatic disease model performance

The rheumatic classification model demonstrated consistent discrimination across cohorts (**Figure 3B**, **Table 5**). In the KPNC held-out test set, AUROC was 0.890 (95% CI, 0.830–0.942) with sensitivity 0.765 and specificity 0.927, and a high NPV of 0.986. Performance was similar in the temporally distinct cohort (AUROC 0.903 [0.831–0.960]). External validation confirmed generalizability, with AUROC 0.944 (0.855–0.994) at SHC and 0.967 (0.955–0.978) at CSMC.

**Table 5.** Rheumatic or non-rheumatic MS model performance.

|  | AUROC | AUPRC | PPV | NPV | Recall | Specificity | F1 |
| --- | --- | --- | --- | --- | --- | --- | --- |
| KPNC held-out (N=950) | 0.890<br>(0.830–0.942) | 0.471<br>(0.353–0.607) | 0.371<br>(0.311–0.440) | 0.986<br>(0.979–0.993) | 0.765<br>(0.647–0.882) | 0.927<br>(0.910–0.943) | 0.500<br>(0.425–0.575) |
| KPNC temporally distinct (N=941) | 0.903<br>(0.831–0.960) | 0.450<br>(0.316–0.611) | 0.265<br>(0.219–0.320) | 0.993<br>(0.987–0.998) | 0.833<br>(0.694–0.944) | 0.908<br>(0.888–0.926) | 0.403<br>(0.337–0.471) |
| SHC (N=1663) | 0.944<br>(0.855–0.994) | 0.541<br>(0.314–0.848) | 0.360<br>(0.258–0.500) | 0.995<br>(0.986–1.000) | 0.900<br>(0.700–1.000) | 0.929<br>(0.893–0.960) | 0.514<br>(0.381–0.667) |
| CSMC (N=234) | 0.967<br>(0.955–0.978) | 0.779<br>(0.714–0.842) | 0.616<br>(0.564–0.672) | 0.984<br>(0.978–0.990) | 0.845<br>(0.784–0.899) | 0.949<br>(0.937–0.959) | 0.712<br>(0.665–0.758) |

Model performance for discriminating rheumatic from non-rheumatic mitral stenosis (MS) among
patients with at least mild MS. AUROC, area under the receiver operating characteristic curve;
AUPRC, area under the precision-recall curve; PPV, positive predictive value; NPV, negative
predictive value; KPNC, Kaiser Permanente Northern California; SHC, Stanford Healthcare; CSMC,
Cedars-Sinai Medical Center

## Discussion

In this study, we developed and validated EchoNet-MS, a novel automated deep learning–based framework that estimates MS severity and classifies MS etiology by incorporating video-based information from multiple echocardiographic views. Our approach demonstrated excellent performance with AUROC ranging from 0.937 to 0.994 for discriminating severe MS, while maintaining favorable NPV and specificity across 4 independent test cohorts.

In recent years, AI has been applied to echocardiography across a range of tasks, including assessment of left ventricular systolic function^25,26^, detection of left ventricular hypertrophy and its various subtypes^27^, detection of valvular regurgitation^8,10,11^, and AS^12,13^. However, to our knowledge, no prior model has been specifically designed to automatically grade MS severity from echocardiography. This likely reflects the relatively low prevalence of MS compared to AS, AR, MR and TR, which has limited the availability of datasets. Leveraging a large, multi-institutional labeled dataset, we developed a first open-source deep learning framework specific for automated MS severity grading and validated its performance across held-out, temporally distinct and external cohorts. More recently, open-source multi-task foundation models have been proposed to support broad phenotyping at scale, demonstrating strong performance across multiple labels including MS^14,15^. However, the number of MS-positive cases in their training datasets was limited (901 studies in PanEcho and 2,097 studies in EchoPrime), whereas our model was trained and validated using a total of 112,236 studies, with 5,239 MS studies in the training dataset. This substantial increase in labeled MS cases likely contributed to the improved performance observed in our study compared with the recent foundation models.

Importantly, our framework also demonstrated strong performance in classifying rheumatic versus non-rheumatic MS, a distinction not addressed by prior foundation models. Rheumatic MS, characterized by commissural fusion and leaflet thickening from the leaflet tips, has been associated with a higher risk of thromboembolic events^1^. In contrast, non-rheumatic (calcific) MS is typically driven by mitral annular calcification and is more commonly observed in elderly patients, individuals with prior chest radiation exposure, or those with end-stage renal disease^2^. Calcific MS has been associated with increased procedural and surgical risk, and percutaneous interventions may be technically more challenging^2,3^. Accordingly, these two etiologies represent distinct pathophysiologic entities with potentially different management strategies. By enabling automated differentiation of these etiologies, our model may support more tailored risk stratification and clinical decision-making in patients with MS.

EchoNet-MS generalized across held-out cohort, temporally distinct cohort, and two external cohorts with differing patient and echocardiogram acquisition characteristics. This breadth of validation reduces the risk that performance reflects idiosyncrasies of a single site or period. Moreover, performance remained robust across subgroups defined by age, sex, and in the presence of AF or other significant valvular diseases. These findings suggest the framework is broadly applicable across typical clinical scenarios. By flagging studies with moderate/severe MS, the system could help reduce the risk of missed significant MS on echocardiography and facilitate more timely intervention. In instances where the model’s prediction disagrees with the physician’s initial interpretation, a secondary review of the echocardiographic videos and reassessment of severity grading could be undertaken, which may further enhance diagnostic precision.

There are a few limitations worth considering. First, the ground truth for MS severity was based on clinical reports rather than a dedicated core lab analysis, which could introduce some variability in the reference standard. However, even with possible heterogeneity in the ground truth label, our model generalized well across institutions. Second, our model was not trained or evaluated in patients with prosthetic mitral valves. Assessment of mitral stenosis in the setting of prosthetic valves is inherently challenging due to valve type–specific normal ranges, acoustic shadowing, and altered hemodynamics. Therefore, the performance and applicability of our model in this population remain uncertain and warrant future investigation. Finally, this study lacks prospective validation; future studies incorporating prospective evaluation, randomized implementation strategies, and integration into clinical workflow^26,28,29^, are needed to determine real-world effectiveness and impact on patient outcomes.

### Conclusions

In summary, we propose EchoNet-MS, an open source, video-based framework that integrates multi-view echocardiographic information to assess the severity and etiology of MS. Validated across four independent cohorts, EchoNet-MS demonstrated robust performance, particularly for detecting clinically significant MS, and could serve as a valuable clinical decision support tool in routine clinical practice.

## Declarations

### Disclosure of Interest

APA reports Research support through grants from the NIH (R01HL173866 and R01AG091005), the FDA (U01FD008704), the AHA, The Permanente Medical Group, Kaiser Permanente Northern California Community Health Programs, Garfield Memorial Fund, Abbott Laboratories, Abiomed, Amarin Pharma, Inc., Bayer Pharma AG, Boehringer Ingelheim, Cordio Medical, Edwards Lifesciences LLC, Esperion Therapeutics, Inc., Merck, and Novartis. Consulting for Bristol Meyer Squibb, Bayer Pharma AG, Corstasis, Merck, Novo Nordisk, Pharmacosmos A/S, Pfizer, scPharma, SQ Innovation, Reprieve Cardiovascular, and VisCardia. DO reports consulting or honoraria for lectures from EchoIQ, Ultromics, Pfizer, InVision, the Korean Society of Echocardiography, and the Japanese Society of Echocardiography.

### Code availability

Our code and model weights are available at https://github.com/echonet/MS.

### Data availability

The dataset of echocardiography videos and reports used to train the model is not available for public sharing, given the restrictions in our institutional review board approval.

### Funding

This work is funded by National Institutes of Health (R00HL157421, R01HL173487 and R01HL173526) and Alexion.

### Ethical approval

This study received approval from the Institutional Review Boards of KPNC, SHC and CSMC. Informed consent was waived due to the study’s use of secondary analysis of existing data.

